# SARS-CoV-2 detection by nasal strips: a superior tool for surveillance of pediatric populations

**DOI:** 10.1101/2020.10.28.20220673

**Authors:** Renee WY Chan, Kate C Chan, Kathy YY Chan, Grace CY Lui, Joseph GS Tsun, Rity YK Wong, Michelle WL Yu, Maggie HT Wang, Paul KS Chan, Hugh Simon Lam, Albert M Li

**Author notes:** **Corresponding Author:** Albert Martin Li, Professor, Address: Department of Paediatrics, 6/F, Lui Chee Woo Clinical Sciences Building, Prince of Wales Hospital, New Territories, Hong Kong.

## Abstract

**Background:** Deep throat saliva (DTS) and pooled nasopharyngeal swab and throat swab (NPSTS) are utilized for viral detection. DTS is challenging for children. Swabbing the respiratory mucosa requires trained personnel and may trigger sneezing and coughing, which generate droplets. A reliable, simple and safe sampling method applicable to a wide age range is required for community-based surveillance.

**Methods:** We introduced nasal strip as an easy and low-risk collection method. Asymptomatic and symptomatic SARS-CoV-2 infected patients (n = 38) were recruited. Nasal epithelial lining fluid (NELF) (n = 43) strip paired with nasal swab (n = 13) were collected by a healthcare worker to compare with NPSTS (n = 21) or DTS (n =22) collected within 24 hours as reference. All samples were subjected to viral RNA quantitation by real-time PCR targeting the nucleoprotein gene.

**Results:** Comparable Ct values were observed between paired nasal strip and nasal swab samples. The agreement between nasal strip samples and NPSTS was 94.44% and 100% for NPSTS positive and negative samples. Higher viral RNA concentration was detected in nasal strips than DTS samples. False-negative results were recorded in six DTS specimens, of which four were from children. Storage at room temperature up to 72 (n = 3) hours did not affect diagnostic yield of nasal strips.

**Conclusions:** Nasal strip is a reliable and non-invasive sampling method for SARS-CoV-2 detection, and viral detection remains stable for at least 72 hours. It can be used as an alternative tool for community-based surveillance.

## Introduction

Many cities have started community surveillance programs for Severe Acute Respiratory Syndrome Coronavirus-2 (SARS-CoV-2) so that local governments can make evidence-based decisions regarding social distancing and school-reopening policies. In the clinical setting, deep throat saliva (DTS) and pooled nasopharyngeal and throat swab (NPSTS) are standardized sample collection methods for early viral detection^1^. However, these methods do not translate effectively to the community setting. DTS can be challenging to obtain from young children as well as the elderly and could invariably reduce the test sensitivity.^1^ In contrast, NPSTS when correctly performed, can yield relatively reliable bio-specimens. However, swabbing respiratory mucosa deep in the nasopharyngeal and throat cavities can be carried out only by trained personnel. Furthermore, the procedure may trigger sneezing and coughing, which poses a risk of disease transmission to people nearby, especially the person collecting the sample. The need for adequate personal protective equipment is thus mandatory.

Tongue^2^, nasal^2,3^, or mid-turbinate swab samples^2^ compared with nasopharyngeal swab specimens have recently been evaluated with respect to sensitivity in detecting SARS-CoV-2 during the course of infection. Though able to achieve comparable sensitivity in viral detection, these methods induce discomfort and therefore limit their use in children and or as a self-administered tool. Recent studies found the presence of SARS-CoV-2 RNA in fecal specimens of COVID-19 patients.^4,5^ Stools with infective signature^6^ and live SARS-CoV-2 have also been observed in some patients.^7^ However the association between viral gene copies detection in fecal specimens and infectivity remains dubious.^8^ In addition, the pooled detection rate of fecal SARS-CoV-2 RNA is modest at 43.7% and 33.7% by patient and number of specimens as a unit count, respectively.^4^ Presence of gastrointestinal symptoms does enhance the detection rate, but this cannot facilitate surveillance work in the community. A non-invasive and reliable self-administered sampling method applicable to a wide age range of patients which does not involve direct interaction between healthcare workers and subjects would be ideal. Stability of the specimens at room temperature is equally important and vital for any large-scale community-based screening programs.

## Methods

### Study design, population and settings

#### Subject Recruitment

Thirty-eight asymptomatic and symptomatic subjects hospitalized with COVID-19 were recruited prospectively by convenience sampling. The disease status was confirmed by two RT-PCR tests targeting different regions of the RdRp gene performed by the local hospital and Public Health Laboratory Service. Adult subjects (n = 20) or guardians in the case of subjects below 18 years old (n = 18) provided informed consent. The study was approved by the Joint Chinese University of Hong Kong – New Territories East Cluster Clinical Research Ethics Committee (CREC: 2020.076 and 2020.442) and took place at the Prince of Wales Hospital.

#### Deep Throat Saliva (DTS) and Pooled Nasopharyngeal and throat swab (NPSTS) collection

For adolescent and adult subjects, they were asked to provide a DTS specimen by making gargling noise so to clear saliva from deep throat and the sample was then spit into a sterile bottle. The sampling was carried out first thing in the morning before tooth brushing and breakfast. A demonstration video produced by the Centre for Health Protection, Hong Kong Special Administrative Region was shown to the subjects beforehand.^9^ For children aged under 5 years and the elderly group, NPSTS was collected using flocked swabs (FLOQSwabs, Copan, Italy) and handled according to standardized protocols by healthcare workers under strict infection control precautions.

#### Nasal epithelial lining fluid (NELF) collection by the nasal strip method

Nasal strips were cut from sheets of Leukosorb medium (Pall Corporation, BSP0669) using a laser cutter (CMA960, Department of Biomedical Engineering, CUHK) to the dimensions of 4mm wide and 40mm long with a marking at 12mm as previously described^10^ for adults and 32mm long for children under 10 year-old. One strip was inserted into each nostril after 100μL of sterile saline was instilled. The strip touching the anterior part of the inferior nasal turbinate was inserted to a depth until the indicator mark was at or close to the base of each naris. Then a 1-minute nose pinch to allow direct contact of the strip against the nasal mucosa was done to facilitate NELF absorption. Both strips were then removed from the nostrils and placed in a dry sterile 2-mL collection tube. The specimens were submerged in viral RNA lysis buffer within 24 hours after collection with a quick vortex or stored at room temperature for an extra 24 or 72 hours to test for their room temperature stability for the detection of SARS-CoV-2. The lysate solution and the strip were then transferred to a Costar Spin-X (CLS9301) and centrifuged at 13,000 rpm for 2 minutes. The flow-through was subjected to viral RNA extraction.

#### Nasal Swab

Briefly dry nasal samples were collected before the collection of nasal strip by gently inserting a flocked swab (FLOQSwabs, Copan, Italy) in a horizontal position. The entire tip of the swab was placed inside the nose, and the side of the swab tip was rubbed with moderate pressure against the wall of the anterior nares region in a large circular path inside the nose. The swab was left in place for 10-15 seconds per nostril while being rotated 5 times and rubbed against the nasal mucosa. The procedure was repeated on the other side with the same swab as described^2^. The swab specimens were then subjected to RNA lysis followed by RNA extraction and viral gene quantification by qPCR.

#### Room temperature (RT) storage of nasal strips

As nasal strip can be self-administered and potentially useful in community surveillance, we further explored sample stability after 24 to 72 hours of storage at RT, so as to assess the feasibility of home sampling followed by postage. Paired nasal strip samples were collected from 6 patients to assess temperature stability. One set of a pair was extracted within 4 hours of collection and the other was randomly assigned to room temperature storage for either 24 or 72 hours before RNA extraction.

#### Viral RNA extraction and quantification

RNA of the nasal strip and nasal swab samples was extracted and eluted in 30μL using QIAamp Viral RNA Mini Kit (Qiagen, Hilden, Germany). SARS-CoV-2 RNA was detected and quantified by real-time reverse-transcriptase polymerase chain reaction (RT-PCR), with primers targeting the N gene of SARS-CoV-2 as described.^11^ For strips and swabs, 8μL of RNA eluate was reverse transcribed with Takara PrimeScript RT cDNA kit according to the manufacturer’s instructions (Takara, Shiga, Japan). The equivalent input quantity for nasal strip or nasal swab was subjected to each 20□μL qPCR reaction including 5μM primers (forward: 5′ TAATCAGACAAGGAACTGATTA and reverse: 5′CGAAGGTGTGACTTCCATG; Thermo Fisher Scientific, Waltham, MA, USA), 2x Master Mix with ROX (Takara, Shiga, Japan). Duplicate reaction was conducted on QuantStudio 12K Flex Real-Time PCR System (Applied Biosystems, Foster City, CA, USA) at the following cycling conditions: 1 cycle at 95.0□°C for 30 seconds and 40 cycles at 95.0□°C for 5□seconds, 60.0□°C for 34 seconds. No template control and a positive control with cDNA synthesized from SARS-CoV-2 infected Vero E6 cells were included in each run.

#### Statistical analysis

McNemar’s test was used to evaluate differences between reference specimens and nasal strip samples. The correlation of Ct values between nasal strip, NPSTS and DTS specimens was examined by Spearman’s correlation test. The Ct values of all specimens from different sampling methods were tested by Wilcoxon signed rank test for matched pairs. Differences were considered to be statistically significant if *p* < 0.05. The analyses were performed with Prism version 8.4.3 for Mac or SPSS version 25 (SPSS Inc, Chicago, IL, USA).

## Results

### Patients demographics

We obtained NELF samples using nasal strips and at least one of the standard methods (DTS or NPSTS) in 38 COVID-19 confirmed patients with a median age of 25 years old. Twenty infected adults (range: 22-74 years old) and eighteen children/adolescents (range: 6 – 17 years old) were recruited of whom ten were asymptomatic.

### Performance of the nasal strip samples

Of the 43 nasal strips collected, 21 and 22 were paired with NPSTS and DTS, respectively. The agreement between nasal strip samples and NPSTS was 94.44% (17/18) and 100% (3/3) for NPSTS for NPSTS positive and negative samples (Table 1). In contrast, the agreement between nasal strip specimens and DTS was 93.33% (14/15) and 14.29% (1/7) for DTS positive and negative samples, respectively (Table 1). Eight discrepant samples were identified (Supplementary Table 1, Fig S1) and of which seven were DTS specimens. Nasal strip outperformed DTS on six occasions, where negative result was reported in the latter. Four of these DTS specimens were collected from pediatric patients (Patients 1 to 4). Nasal strip samples were tested negative on two occasions when the reference test revealed Ct values of 35 and 28.92. Spearman’s test demonstrated significant correlation between NPSTS and nasal strip (*p* = 0.0003) and DTS and nasal strip (*p* = 0.0106) (Figure 1A). Bland-Altman plots indicated that the nasal strip give consistent and comparable measurements versus the NPSTS (Figure 1B) and DTS (Figure 1C). Further analysis by Wilcoxon signed rank test revealed that nasal strip and NPSTS gave similar Ct values (Figure 1D, *p* = 0.76) while a lower Ct was detected in nasal strip compared to paired DTS (Figure 1E, *p* = 0.016).

**Table 1.** Number of tested samples and performance of the each collection methods.

| Reference |  | Nasal strip |  |  |  |
| --- | --- | --- | --- | --- | --- |
|  |  | Positive | Negative | Total | Agreement (%) |
| NPSTS<br>(n = 21) | Positive | 17 | 1 | 18 | 94.44 |
|  | Negative | 0 | 3 | 3 | 100.00 |
|  |  | Value (95% CI) |  |  |  |
| Accuracy |  | 95.2 (76.18-99.88) |  |  |  |
| DTS<br>(n = 22) | Positive | 14 | 1 | 15 | 93.33 |
|  | Negative | 6 | 1 | 7 | 14.29 |
|  |  | Value (95% CI) |  |  |  |
| Accuracy |  | 68.18 (45.13-86.14) |  |  |  |

No significant difference in the detection rate was observed between NPSTS (McNemar’s test $p = 1.000$ ) or DTS ( $p = 0.125$ ) for the qPCR test (NPSTS and DTS, $p = 0.289$ ). CT value equal or below 35 is defined as positive.

**Figure 1.**
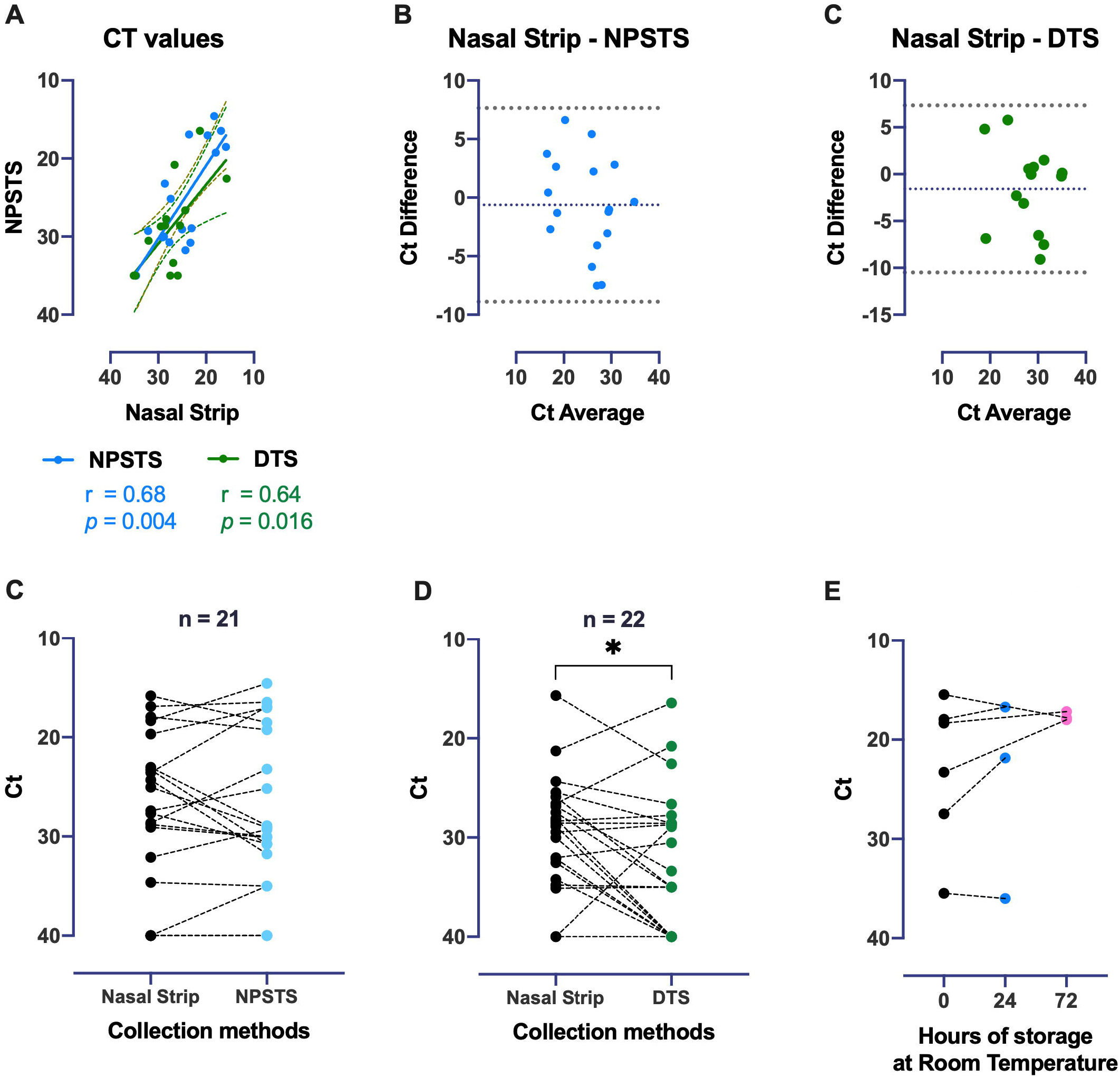
Correlation, agreement and comparison of the cycle threshold (Ct) values from nasal strip, NPSTS and DTS, and the stability of nasal strip sample at room temperature. The correlation coefficients of NPSTS and DTS are superimposed on the panel with trend lines estimated with the use of simple linear regression (Panel A). Plot shows the available Ct values of 31 samples which had positive test result from both tests. Data on three samples with negative result in both nasal strip and NPSTS, one sample with negative result in nasal strip but a positive result in NPSTS (Ct value = 35), one sample with negative result in both nasal strip and DTS, one sample with negative result from nasal strip but positive result in DTS, and six samples with positive result in nasal strip but negative result DTS were excluded from the Spearman correlation analysis. Bland-Altman Plots indicate agreement of nasal strip versus NPSTS (Panel B) and DTS (Panel C), respectively. The difference between the two measurements are plotted against their average Ct values. Almost all observations are located within 2 standard deviations of the mean difference, and no bias is shown. The plots show that the nasal strip give consistent and comparable measurements versus the NPSTS and DTS. SARS-CoV-2 RNA load in nasal strip and NPSTS (n = 21) (Panel D) and DTS (n= 22) (Panel E). Samples were obtained from 36 in-patients who had a diagnosis of COVID-19. Panel A shows SARS-CoV-2 RNA Ct in the nasal strip and NPSTS; panel B shows SARS-CoV-2 RNA Ct in the nasal strip and DTS. The lines indicate samples from the same patient obtained within 24 hours. Negative result is arbitrarily set as Ct = 40 and results were compared with the use of a Wilcoxon signed-rank test (*p* < 0.05). Panel F shows the stability of nasal strip sample for the detection of SARS-CoV-2 (n = 6). Comparison of Ct upon 24 (blue) and 72 (pink) hours RT storage from nasal strips directly lysed after sample collection.

### Comparison between nasal strip and nasal swab samples

Of the 43 nasal strips collected, 13 were paired with a nasal swab sample obtained concurrently by a healthcare worker. A significant correlation was found between Ct values from the nasal strip and nasal swab specimens (Figure S2A). Though nasal swab missed two positive cases detected by nasal strip and nasal strip missed one positive case detected by nasal swab, there was no significant difference detected between Ct values of the 13 paired samples (Figure S2B).

### Validity of nasal strip samples after prolonged room temperature storage

Finally, we collected nasal strip pairs from six patients to determine viral RNA stability over time, viral RNA remained detectable after 24- and 72-hour storage in room temperature (Figure 1F).

## Discussion

We introduced nasal strip as a non-invasive and user-friendly sampling tool for SARS-CoV-2 detection. Both asymptomatic and symptomatic laboratory confirmed SARS-CoV-2 infected patients (n = 38) were recruited to validate this method. The high correlation of nasal strip samples with the standard sampling methods and its high agreement is likely the result of steady NELF absorption with the strip in close contact with the nasal mucosa which reduces sample variability. This study also indicated the possible insensitivity of DTS, particularly in pediatric patients who are less able to provide DTS with consistent quality (Supplementary Table 1) and how nasal strip would be a superior tool for surveillance of paediatric populations. Nasal strip is also a better collection method than NPSTS as it is less traumatic and irritating. The application of nasal strip reduces the risk of any sneezes and coughs and therefore lessens the risk of virus transmission. Nasal strip is a more comfortable and easier to apply sampling method compared with the other available standard sampling tools. Repeat nasal strip sampling as part of a community-based surveillance program is feasible in children and adults and likely to succeed as a result of its non-invasive nature (Video 1).

Nasal swabbing is considered relatively easy to perform by healthcare workers. It was evaluated as an alternative sample collection method in this study and we found good agreement in Ct values from nasal strip and nasal swab specimens. However, nasal swab, as self- or parent-assisted application, may not be possible for children and it can be a potentially high-risk procedure without supervision. The need for healthcare or trained personnel to carry out a procedure is a source of testing bottlenecks and presents a chance of disease transmission.

Compared with NPSTS, nasal strip sampling achieved an accuracy of 95.2% (Table 1). This is comparable if not superior to other sampling methods reported in the literature, including self-administered tongue, lower- and mid-nasal specimens.^2^ Apart from a good accuracy, we assessed the validity of the nasal strip samples after prolonged storage at room temperature so as to mimic the duration needed to post the specimens to the laboratory. The validity of the sample stability after prolonged room-temperature storage was not assessed in previous studies, albeit an important criterion if a sampling method is adopted for community-based testing purposes. Our findings suggest that nasal strip would provide at least consistent qualitative results (positive or negative), as long as the Ct value is within the range of an inferred infectivity^12^. This would be sufficient to identify potentially infectious individuals and susceptible contacts for further management and quarantine.

There are several limitations in this study. This prospective study presents the cross-sectional data performed in a single hospital and limited to the performance comparisons with the results of NPSTS and DTS, which are not perfect standard tests. The clinical sample pairs (n = 6) that underwent 24- to 72-hour RT storage remained stable in terms of viral detection. However, the involvement of protease and RNase activity of individual subjects and its contribution to sample stability was not fully elucidated. Moreover, we did not evaluate the quality of the sample in a genuine home to laboratory scenario. If extended sample transportation period is necessary, stabilization buffer e.g. RNAlater might be needed. Lastly, the current method provided detection of SARS-CoV-2 at the gene level but no information was obtained regarding the infectious titer.

## Conclusion

Our nasal strip collection method serves as an excellent sampling method with comparable performance with NPSTS, DTS and nasal swab specimens in identifying subjects infected with SARS-CoV-2. This reliable, non-invasive, self-administered method with its extended sample stability makes it uniquely suited for repeated sampling and large-scale community study, especially for pediatric population.

## Data Availability

Not applicable

## Acknowledgements

We would like to acknowledge Prof Aaron HP Ho, Prof Megan YP Ho and Miss Yuan-yuan Wei (Department of Biomedical Engineering, Faculty of Engineering, The Chinese University of Hong Kong) who tailor-cut the nasal strips for this study and Dr KP Tao and Ms Fiona Cheng (Department of Pediatrics, The Chinese University of Hong Kong) for his technical support in molecular biology and her assistance in preparing all the sampling kits, respectively. We would like to offer our special thanks to Mr. Ernie Lee, Mr. Johnson Chan and Mr. Reneson Chan in contributing consecutive NELF samples during the testing stage and all the subjects who agreed to participate in this study. We thank Prof Ellis KL Hon (Department of Pediatrics, The Chinese University of Hong Kong) for his continuous encouragement to the research team.

## Authors contributions

Conception, experimental design, drafting the manuscript and interpretation: RWYC, KYYC, HSL, AML; Experimental design and analysis: RWYC, KYYC, GCYL, MHTW, PKSC; Patient recruitment: RWYC, KCCC, GCYL, JGST, MY, RYKW, AML; Acquisition of data: RWYC, KCCC, KYYC, JGST, AML; Nasal strip protocol optimization: RWYC & JGST. All authors reviewed and approved the final manuscript.

## Legend

**Video 1**. Illustration of the self-administering procedures to collect the NELF sample using nasal strip in child.

https://youtu.be/qCplX-ounPA

