## Supplementary Appendix for "SARS-CoV-2 detection by nasal strips: a superior tool for surveillance of pediatric populations": medRxiv Supplementary.docx

**Table S1.** Details of discrepant samples between reference and nasal strip test.

**Fig S1. Longitudinal Ct values detected by NPSTS and DTS from the eight subjects with discrepant result with nasal strip as listed in table S1.** Vertical dotted line denotes the time of nasal strip sampling while the red dot represents the Ct value of the nasal strip. Blue square and black dot represent the Ct value of NPSTS and DTS sample collected along the disease course, respectively.^1^ Negative result is arbitrarily set as Ct = 40. We confirmed absence of viral nucleoprotein gene detection in the nasal strip samples of patient 7 and patient 8 was not due to the RNA extraction failure, human housekeeping gene (beta-actin) expression was detected in these two samples with a Ct value of 26.33 and 25.21, respectively.

**Fig S2. Comparison of paired nasal strip and swab specimens**. Panel A shows the scatter plot of the Ct values of 9 samples which had positive test result from both tests. Negative tests results were excluded. The Spearman correlation coefficient between nasal swab and nasal strip is r = 0.88 with *p* = 0.0031. Panel B shows SARS-CoV‐2 RNA Ct values in the nasal strip and nasal swab, respectively. The lines connect samples from the same patient obtained concurrently. Negative result is arbitrarily set as Ct = 40. Results were compared with the use of a Wilcoxon signed‐rank test (*p =* 0.22).
