## Supplementary figures and images for "SARS-CoV-2 detection by nasal strips: a superior tool for surveillance of pediatric populations"

### medRvix_Table S1.tif

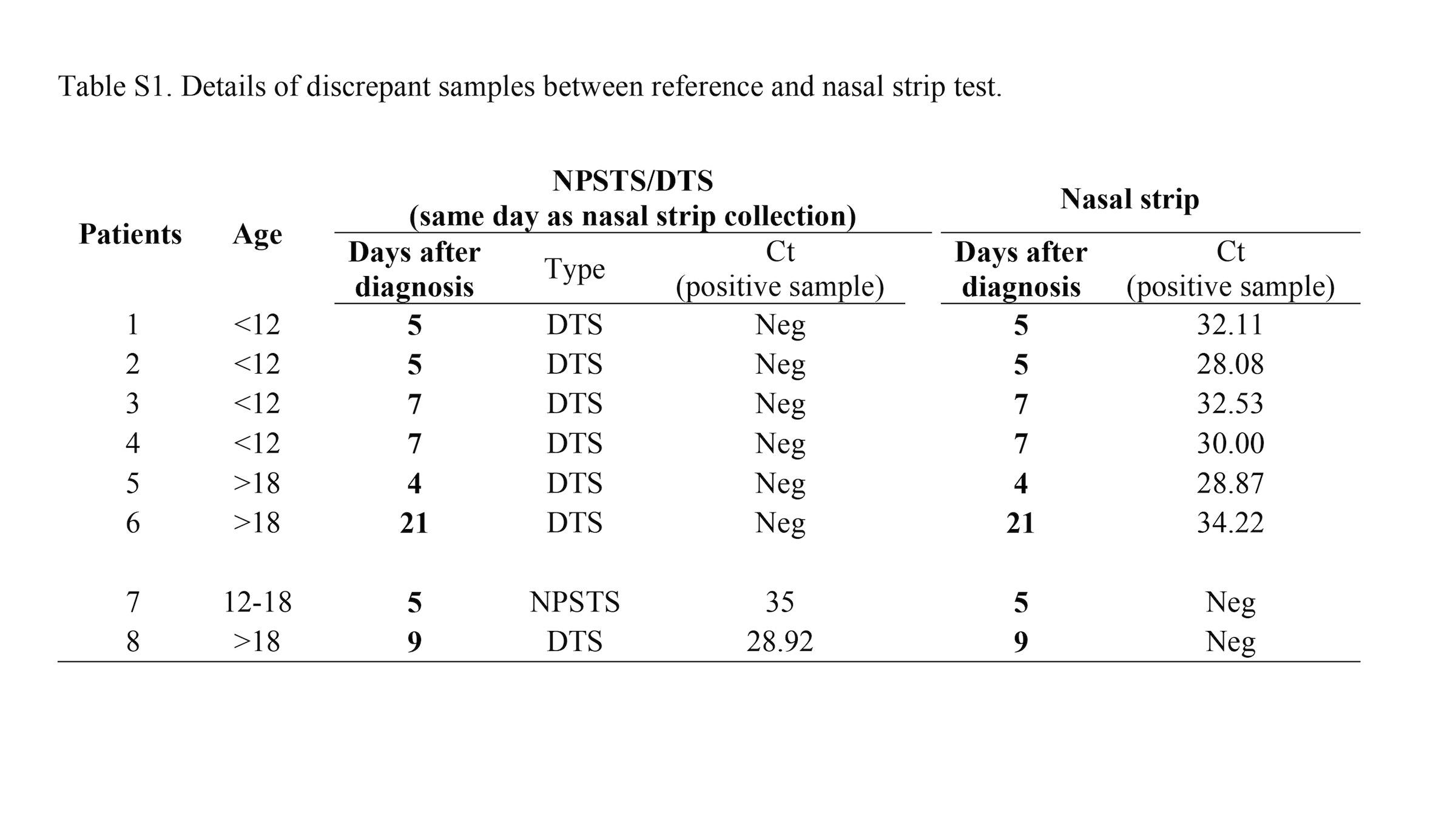

### medRxiv_FigS1.tiff

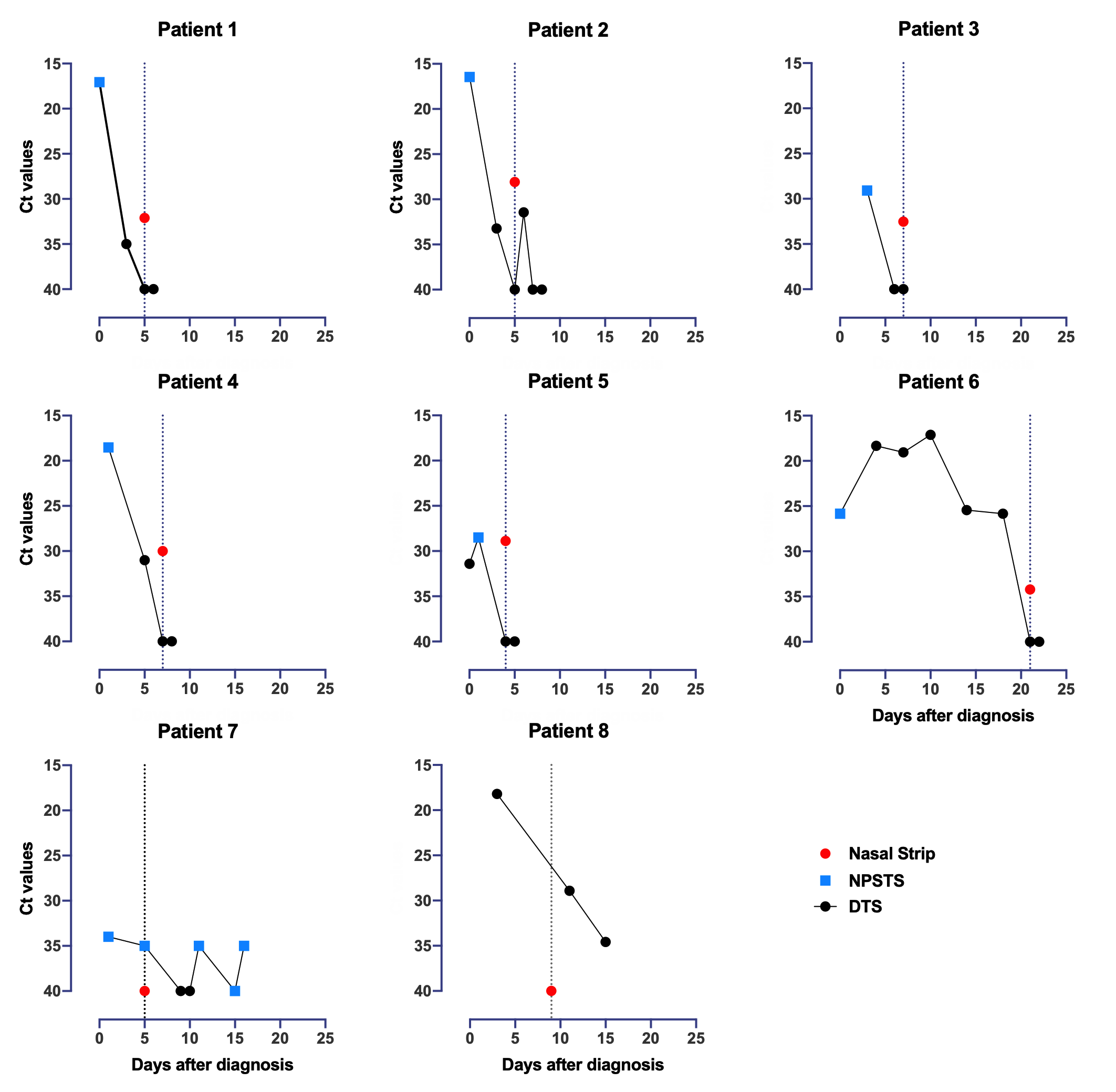

### medRxiv_FigS2.tiff

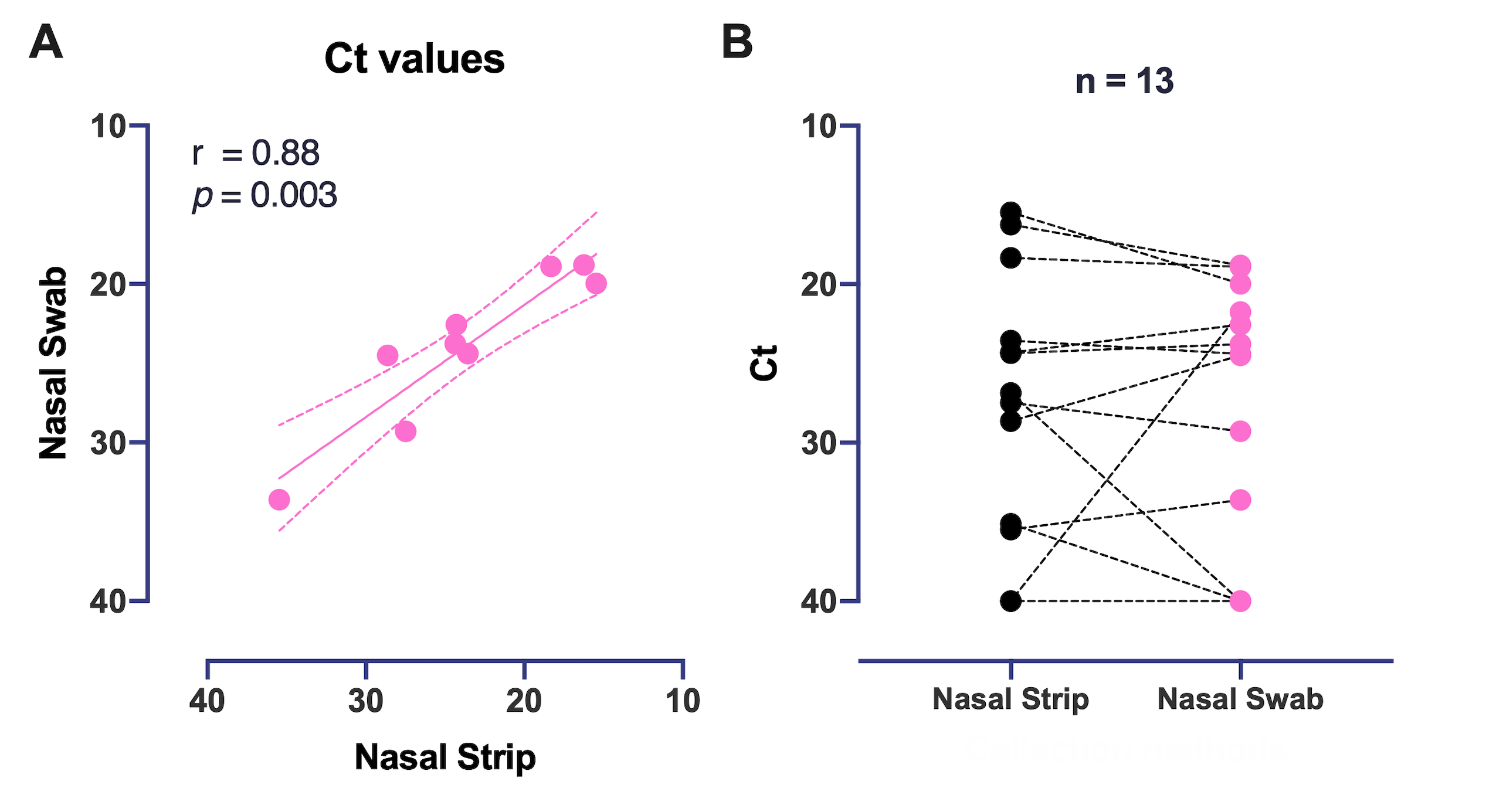
